# Breastfeeding of Infants Born to Mothers with COVID-19: A Rapid Review

**DOI:** 10.1101/2020.04.13.20064378

**Authors:** Nan Yang, Siyi Che, Jingyi Zhang, Xia Wang, Yuyi Tang, Jianjian Wang, Liping Huang, Chenglin Wang, Hairong Zhang, Muna Baskota, Yanfang Ma, Qi Zhou, Xufei Luo, Shu Yang, Xixi Feng, Weiguo Li, Toshio Fukuoka, Hyeong Sik Ahn, Myeong Soo Lee, Zhengxiu Luo, Enmei Liu, Yaolong Chen, on behalf of COVID-19 evidence and recommendations working group

## Abstract

**Background:** Existing recommendations on whether mothers with COVID-19 should continue breastfeeding are still conflicting. We aimed to conduct a rapid review of mother-to-child transmission of COVID-19 during breastfeeding.

**Methods:** We systematically searched Medline, Embase, Web of Science, Cochrane library, China Biology Medicine disc, China National Knowledge Infrastructure, Wanfang, and preprint articles up to March 2020. We included studies relevant to transmission through milk and respiratory droplets during breastfeeding of mothers with COVID-19, SARS, MERS and influenza. Two reviewers independently screened studies for eligibility, extracted data, assessed risk of bias and used GRADE to assess certainty of evidence.

**Results:** A total of 4481 records were identified in our literature search. Six studies (five case reports and one case series) involving 58 mothers (16 mothers with COVID-19, 42 mothers with influenza) and their infants proved eligible. Five case reports showed that the viral nucleic acid tests for all thirteen collected samples of breast milk from mothers with COVID-19 were negative. A case series of 42 influenza infected postpartum mothers taking precautions (hand hygiene and wearing masks) before breastfeeding showed that no neonates were infected with influenza during one-month of follow-up.

**Conclusions:** The current evidence indicates that SARS-CoV-2 viral nucleic acid has not been detected in breast milk. The benefits of breastfeeding may outweigh the risk of SARS-CoV-2 infection in infants. Mothers with COVID-19 should take appropriate precautions to reduce the risk of transmission via droplets and close contact during breastfeeding.

## Background

In December 2019, a pneumonia caused by a previously unknown coronavirus emerged in Wuhan, Hubei Province, China. During the subsequent weeks and months, the disease, later named COVID-19, spread rapidly nationwide and globally, and was declared a global pandemic by the World Health Organization.

Existing studies have confirmed that all people are susceptible to this novel coronavirus (SARS-CoV-2) (1). Cases of COVID-19 among pregnant and lactating women have also been confirmed (2). Chinese guidelines recommend suspending breastfeeding if the mother is suspected or confirmed with COVID-19 (3). The Centers for Disease Control and Prevention of the USA have published recommendations for mothers with COVID-19 and their family members and healthcare providers on whether and how to start or continue breastfeeding (4). However, none of the above recommendations provide relevant supporting evidence.

So far, there is no consensus reached on whether mothers with COVID-19 should continue breastfeeding. This question is also listed as a priority in the *Rapid Advice Guidelines for Management of Children with COVID-19* (5), currently under development. We thus conducted this rapid review on studies of mother-to-child transmission of COVID-19 during breastfeeding to provide evidence support for clinical decision-making.

## Methods

### Search strategy

Considering of the few search results on COVID-19 based on strategy preliminary search by the guidance panel, the rapid review also searched studies on breastfeeding for Severe Acute Respiratory Syndrome (SARS), Middle East Respiratory Syndrome (MERS) and influenza. Two reviewers (N Yang and S Che) adopted the following terms by consensus: “breast feeding”, “lactation”, “milk”, “COVID-19”, “novel coronavirus”, “2019-novel coronavirus”, “Novel CoV”, “SARS-CoV-2”, “2019-CoV”, “Middle East Respiratory Syndrome”, “MERS”, “Severe Acute Respiratory Syndrome”, “SARS”, “influenza” and “flu” and their derivatives (Full search strategies are presented in Supplementary Appendix 1). Two groups (N Yang and J Wang, N Yang and H Zhang) carried out the search independently in the following electronic databases: Medline (via PubMed), Embase, Web of Science, the Cochrane library, China Biology Medicine disc (CBM), China National Knowledge Infrastructure (CNKI), and Wanfang Data. All databases were searched from their inception until March 31, 2020. Two authors (N Yang and S Che) also searched the following websites for relevant publications: World Health Organization (WHO), the National Health Commission of the People’s Republic of China, Google Scholar, BioRxiv, SSRN, and MedRxiv. We also scanned published online articles on COVID-19 in selected major medical journals (*Journal of the American Medical Association, The Lancet, New England Journal of Medicine* and their sister journals) and journals related to maternal and pediatric health (*Pediatrics, Journal of Pediatrics and the Chinese Journal of Applied Clinical Pediatrics, Chinese Journal of Pediatrics, Chinese Journal of Neonatology, Chinese Journal of Pediatrics, Chinese Journal of Perinatal Medicine, Chinese Journal of Obstetrics and Gynecology*) to identify potentially eligible studies published on March 31, 2020. The references of included studies were also scanned to identify other eligible studies.

### Inclusion and exclusion criteria

We included studies relevant to transmission through milk and respiratory droplets during breastfeeding of mothers with COVID-19, SARS, MERS and influenza. We did not restrict the studies based on mode of delivery (vaginal or cesarean section), or protective measures during breastfeeding. We excluded duplicates, conference abstracts, comments and letters, studies published in languages other than English or Chinese, and studies where we could not access the full text.

### Study selection

Three groups of two to three reviewers (M Baskota and J Zhang; X Wang and N Yang; Y Tang, L Huang and C Wang) independently screened the titles and abstracts to identify potentially eligible studies, and then read the full‐texts of the potentially eligible studies decide about final inclusion. We reported reasons for exclusion of the ineligible studies. We recorded the selection process in a PRISMA flow diagram and a table showing the characteristics of excluded studies (6). Disagreements were solved through discussion with another researcher (S Che or E Liu).

### Data extraction

Two reviewers (N Yang and S Che) independently extracted the following data from the included studies using a pre-defined extraction sheet: 1) basic information (year of publication, first author, affiliation, year, study type); 2) number of women, age, disease or virus, mode of delivery; 3) number of infants, gender, birth weight, gestational age, APGAR score, signs of infection, results of RT-PCR tests of breast milk or throat swab, and 4) protective measures used during breastfeeding (wearing a surgical mask; hygiene measures taken before close contact with the infant). We conducted a pilot test before the full extraction to ensure that both reviewers agreed with the extraction criteria and process. Disagreements were solved through discussion with a third researcher (Y Chen).

### Risk of bias assessment

Two reviewers (N Yang and S Che) assessed the quality of the included studies independently and cross-checked the results. Disagreements were solved through discussion with a third researcher (Y Chen). We used the Cochrane bias risk assessment tool for randomized controlled trials (7), Newcastle-Ottawa scale (NOS) for case control studies (8), the Institute of Health Economics’ case series quality appraisal tool (IHE) for case series (9), and the Joanna Briggs Institute’(JBI) case report quality appraisal tool for case reports (10) to assess the risk of bias.

### Data synthesis

Comparable data from studies with one outcome were pooled using Review Manager version 5.3 (11). For dichotomous outcomes we calculated the risk ratios (RR) and the corresponding 95% confidence intervals (CI) and *P* values. For continuous outcomes, we calculated the mean difference (MD) and its corresponding 95%CI when means and standard deviations (SD) were reported. When effect sizes could not be pooled, we reported the study findings narratively.

### Quality of the evidence assessment

We assessed the certainty of the estimates of certainty of evidence using the Grading of Recommendations Assessment, Development and Evaluation (GRADE) approach (12). We considered potential limitations in risk of bias, inconsistency, imprecision, indirectness and publication bias (13-16).

As COVID-19 is a public health emergency of international concern and the situation is evolving rapidly, our study was not registered in order to speed up the process (17).

## Results

### Study selection and characteristics

A total of 4481 records were identified by the literature search (*Figure 1*). After screening the titles and abstracts, we retrieved 44 articles for full-text screening. Six articles were finally included for data extraction (*Table 1*): five case reports (2, 18-21) from China published in 2020 on mothers with COVID-19 and their infants, and one case series (22) from the US published in 2013 reporting the effect of precautions for breastfeeding mothers infected with influenza. Because of effect sizes could not be pooled, we reported the study findings narratively.

**Table 1.** Clinical Characteristics of the Mothers with Influenza or SARS-CoV-2 and Their Infants

| Study (Author year) | Chen<br>2020(2) | Zhang<br>2020(18) | Lei<br>2020(19) | Dong<br>2020(20) | Wang<br>2020(21) | Cantey 2013(22) |  |
| --- | --- | --- | --- | --- | --- | --- | --- |
| Period of observation | 2020 | 2020 | 2020 | 2020 | 2020 | 2003-2005 | 2009-2010 |
| Study site | Wuhan,<br>China | Haikou,<br>China | Wuhan,<br>China | Wuhan,<br>China | Wuhan,<br>China | Dallas, USA |  |
| Mothers |  |  |  |  |  |  |  |
| Number of mothers | 9 | 1 | 4 | 1 | 1 | 23 | 19 |
| Median age(years) | 30 | - | 32 | 29 | 34 | 26 | 26 |
| Type of infection |  |  |  |  |  |  |  |
| Influenza | 1 | 0 | 0 | 0 | 0 | 23 | 19 |
| SARS-CoV-2 | 9 | 1 | 4 | 1 | 1 | 0 | 0 |
| Mode of delivery |  |  |  |  |  |  |  |
| Vaginal | 0 | 1 | 1 | 0 | 0 | 14 | 14 |
| Cesarean section | 9 | 0 | 3 | 1 | 1 | 9 | 5 |
| Infants |  |  |  |  |  |  |  |
| Number of infants | 9 | 1 | 4 | 1 | 1 | 23 | 19 |
| Girl | - | 1 | - | 1 | 0 | 9 | 10 |
| Boy | - | 0 | - | 0 | 1 | 14 | 9 |
| Median birth weight (g) | 2789 | - | 2803 | 3120 | 3205 | 3123 | 3333 |
| Median gestational age<br>(weeks), | 37 | - | 36 | 38 | 40 | 38 | 40 |
| Age | Neonate | 3 m 19 d | Neonate | Neonate | Neonate | Neonate | Neonate |
| Median APGAR score |  |  |  |  |  |  |  |
| 1 min | 9 | - | 9 | 9 | 8 | 8 | 9 |
| 5 min | 10 | - | 10 | 10 | 9 | 9 | 9 |
| Breast milk RT-PCR |  |  |  |  |  |  |  |
| Positive | 0 | 0 | 0 | 0 | 0 | - | - |
| Negative | 6 | 1 | 4 | 1 | 1 | - | - |
| Breastfeeding |  |  |  |  |  |  |  |
| Wearing a surgical mask | - | 0 | - | - | - | 0 | 19 |
| Hygiene measure before<br>close contact with infant | - | 0 | - | - | - | 23 | 19 |
| Signs of infection | 0 | 1 | 0 | 0 | 0 | 0 | 0<sup>a</sup> |
| Throat swab RT-PCR |  |  |  |  |  |  |  |
| Positive | 0 | 0 | 0 | 0 | 0 | - | 0 |
| Negative | 6 | 1 | 4 | 1 | 1 | - | 1<sup>b</sup> |

-: Not reported; a: Two infants lost to follow-up; b: One infant with fever (source not identified); influenza rapid test and polymerase chain reaction test negative

**Figure 1.**
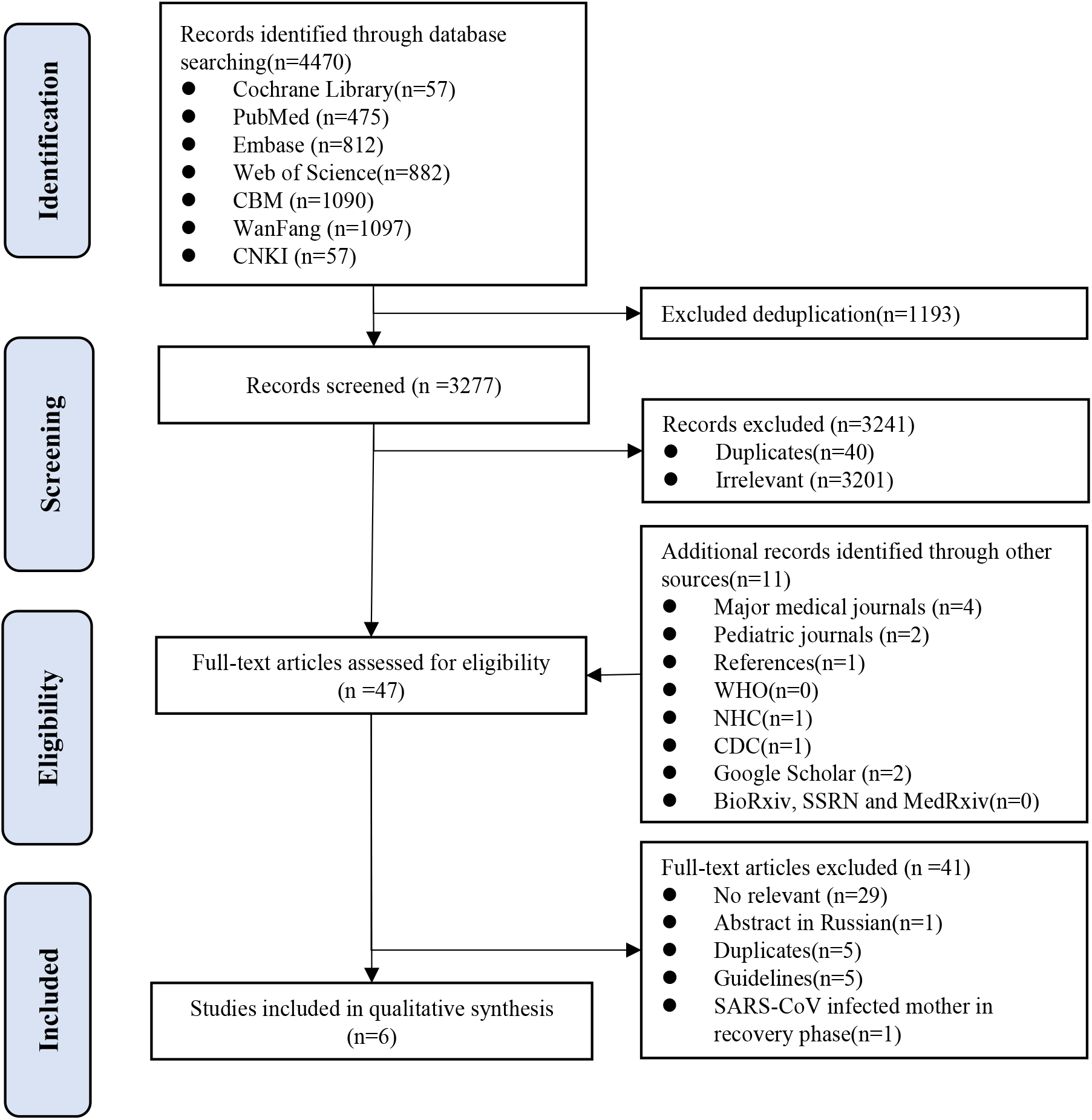
PRISMA flow diagram of the study selection

### Quality assessment

The quality of the included case series study was high: the study met 15 of the 20 items of the IHE evaluation tool. The methodological qualities of the included five case reports studies were also high, all studies meeting 8 of the 8 items of the JBI quality appraisal tool.

### Breastfeeding

Because of no comparable data could be pooled, we reported the study findings narratively. The five case reports (2, 18-21) included sixteen breastfeeding mothers with COVID-19. The first case report showed that neonates of nine mothers with COVID-19 were isolated immediately after delivery and fed with formula. All samples of breast milk from six mothers and throat swab from their infants showed negative nucleic acid test results for SARS-CoV-2 (2). The second case report described a familial cluster of SARS-CoV-2 infection. The nucleic acid tests of throat swab samples from the father, the mother and her three-month-old baby were all positive for SARS-CoV-2, while the breast milk turned out negative (18). The mother did not take any precautions while breastfeeding. The third case report included nine pregnant women infected with SARS-CoV-2, among whom four delivered during the study. The nucleic acid tests of breast milk samples from these four mothers and nasopharyngeal swab samples from their neonates were all negative for SARS-CoV-2 (19). The fourth case report included a mother infected with SARS-CoV-2. The PCR test of breast milk sample was negative six days after delivery, while the nasopharyngeal swab of the mother remained positive eight days after delivery (20). The last case report represented a neonate, who was delivered from a mother with COVID-19 by cesarean section under precautions. The pharyngeal swab specimen from the infant (collected immediately 36 hours after birth) was positive for SARS-CoV-2, and the breast milk of the mother (36 hours after birth) was negative (21).

The case series on influenza included 42 postpartum women, who had a positive influenza test within eight days preceding delivery or in the postpartum period before discharge, during the influenza seasons of 2003 to 2005, and 2009 to 2010. None of the newborns were isolated from their mothers, and all were breastfed. The mothers and infants were followed-up for one month after delivery. During the season of 2003 to 2005, 23 infected mothers washed their hands thoroughly before breastfeeding, but did not wear masks. The 19 mothers observed during the 2009-2010 season both washed hands and wore masks during the. In the end of follow-up, two neonates were lost to follow-up, one neonate had fever with negative nucleic acid test result, and the other 39 neonates did not report any influenza-like symptoms (22).

### Quality of the evidence assessment

Because of the limitations in the study results, we could not perform the assessment of quality of evidence.

## Discussion

This rapid review included five case reports with data on breast milk samples from thirteen mothers with COVID-19 (2, 18-21). The test results for SARS-CoV-2 nucleic acid were all negative. Another case series included 42 influenza infected postpartum mothers taking precautions (hand hygiene and wearing masks) while breastfeeding. During one-month of follow-up, no neonates were infected with influenza (22).

Breast milk is considered by the WHO and other authorities to be the most suitable and nutritious food for babies (23). Multiple studies have investigated the short-term and long-term benefits of breastfeeding from both the infants’ and their mothers’ aspects. Breast milk contains lactoferrin, immunoglobulins, free fatty acids, polyunsaturated fatty acids and growth factors, and it enhances various biological activities such as antimicrobial and immunomodulatory activity. Compared with formula feeding, breastfeeding can promote the development of the infants’ nervous system, and reduce the risk of allergies and infectious diseases. Moreover, breastfeeding can also reduce the risk of breast and ovarian cancer for mothers (24).

Although benefits of breastfeeding are obvious, contraindications should be considered in terms of infectious diseases. The US Centers for Disease Control and Prevention (CDC) (25) advises against breastfeeding if the mother is infected with human immunodeficiency virus (HIV) (26), while the WHO recommends HIV-infected mothers who are fully supported for antiretroviral therapy should breastfeed (27). The US CDC also recommends that mothers infected with human type I or type II T-cell lymphovirus, or Ebola virus (28) should not breastfeed or express breast milk. These viruses have been confirmed to be transmitted via breast milk, and once infected, they can induce long-term detrimental effects on children’s growth and development, and even increase infant mortality. Also if the mother is infected with untreated brucellosis or active herpes simplex virus (29), the US CDC highly recommends suspending breastfeeding or expressing breast milk until the infection gets well treated and controlled. In cases of untreated active tuberculosis or chickenpox during active period, the mother should stop breastfeeding, but expresses breast milk instead (30).

As for influenza, another respiratory contagious disease that also transmits via droplets and close contact, the US CDC has after the first epidemic in the US in 2009 revised the management guidelines for pregnant or lactating mothers infected with H1N1. The first edition of the 2009 guideline recommended that isolation should be implemented to avoid close contact between newborns and H1N1 infected mothers with abnormal body temperature. Expressing breast milk to be fed by healthy caregivers should be encouraged. Subsequently, in consideration of the viability of isolation and the failure of lactation, the guideline was revised to temporarily isolate newborns from H1N1-infected mothers. Isolation could be performed either in separate rooms, or rooming-in with a physical isolation between the mother and child. Expressing breast milk was still recommended. The latest version of the guideline from 2019 recommends that lactating women with influenza can continue breastfeeding or express breast milk. Another CDC guideline targeting hospitalized postpartum women and their newborns recommended that mothers should be temporarily isolated from their newborns to reduce the risk of influenza virus transmission to newborns. Isolation should be decided after discussing the risk of mother-to-child transmission and the benefits of breastfeeding in coordination with the mother’s wishes. If the mother and her baby are temporarily isolated, breast milk should be expressed and fed by healthy caregivers. If the mother and her baby are in the same room, appropriate physical separation should be ensured. Breast milk can be expressed and fed by healthy caregivers. If there is no other caregiver present, the mother can breastfeed using the necessary precautions (such as wearing a mask and taking care of hand hygiene) (31). However, a survey of influenza infection control measures in US maternal and infant healthcare centers showed that the guidelines were not consistently followed: during the 2009 H1N1 epidemic season 58% of hospitals restricted breastfeeding of mothers with influenza-like illness in 2009, while only 31% of hospitals did so in 2010 (32).

COVID-19 is relatively contagious with an estimated basic reproduction number R_0_ of 3.49 (95% CI: 3.42 to 3.58) based on 27,100 confirmed cases from January 17 to February 8, 2020 in Hubei province without intervention (33), which is similar to SARS (R_0_ 2.2-3.6) (34), but clearly higher than Influenza A H1N1 (R_0_ 1.4-1.6) (35). But according to the five case reports (2, 18-21) found by our review the nucleic acid test results for SARS-CoV-2 of breast milk samples from thirteen mothers with COVID-19 were all negative. No studies have reported the existence of virus nucleic acid in breast milk from mothers infected with SARS, MERS or influenza. Based on the considerations above, we believe that SARS-CoV-2 is unlikely to be transmissible via breast milk, but the quality of evidence remains low. As SARS-CoV-2 is transmitted via close contact and droplets, transmission from mother to infant may be possible while breastfeeding. A case series indicated that influenza infected breastfeeding women who washed hands thoroughly did not transmit to their children during a one-month follow-up, regardless of if the mothers wore a mask or not (22). A systematic review conducted in 2016 also revealed that wearing masks and hand hygiene can notably decrease the possibility of respiratory virus transmission (wearing masks: OR 0.32, 95% CI, 0.26 to 0.39; hand hygiene: OR 0.54, 95% CI, 0.44 to 0.67) (36). Therefore, we believe that by taking effective precautions, the risk of transmission while breastfeeding can be reduced.

The effective precautions we recommend can reduce the risk but not avoid the transmission of SARS-CoV-2. Existing studies have however reported that less than 1% of confirmed COVID-19 patients are children under ten years old (37). Chinese Center for Disease Control and Prevention has identified 731 cases of COVID-19 under the age of eighteen years (including 21 sever cases), and 86 cases under the age of one year until February 8th, 2020 (38).

For mothers with severe COVID-19, who need to be treated individually in intensive care units, breastfeeding is difficult to arrange. For children, underlying diseases underlying diseases such as congenital heart disease, immunodeficiency disease and bronchopulmonary dysplasia may increase the risk of severe respiratory infection (39). Under these circumstances, expressing breast milk should be encouraged. If using a breast pump, strict washing and sterilization procedure should be followed. Nevertheless, given the accessibility of expressing breast milk after the outbreak of COVID-19, we agree with the recommendation of US CDC that whether and how to start or continue breastfeeding should be determined by the mother in coordination with her family and health care practitioners.

This study is the first rapid review of breastfeeding for mothers with COVID-19. Following limitations exist: 1) the relevant evidence included is scant and the quality of evidence is low; 2) the evidence is based on small samples of breast milk that were taken only once from each mother, so the presence of virus in breast milk over time cannot be assessed; 3) because of the small number of cases reported, the understanding about COVID-19 in children is not yet comprehensive enough. Therefore, we believe longitudinal studies testing series of breast milk samples from lactating women with different courses and degrees of severity of the disease should be encouraged. Future studies to support comprehensive understanding of the epidemiology, clinical characteristics, the proportion of severe cases, risk factors, and prognosis in children with COVID-19 are also needed. Moreover, cohort studies of breastfeeding mothers with COVID-19 taking effective precaution measures will definitely provide better evidence to inform this issue.

## Conclusion

There is currently no evidence of detected viral nucleic acid in breast milk of mothers infected with SARS-CoV-2. Taking appropriate precautions can reduce the likelihood of transmission via droplets and close contact during breastfeeding. Furthermore, most of the cases in children with COVID-19 have been mild or atypical. The benefits of breastfeeding are thus likely to outweigh the risk of COVID-19 infection in infants.

## Data Availability

A total of 4481 records were identified in our literature search. Six studies (five case reports and one case series) involving 58 mothers (16 mothers with COVID-19, 42 mothers with influenza) and their infants proved eligible. Five case reports showed that the viral nucleic acid tests for all thirteen collected samples of breast milk from mothers with COVID-19 were negative. A case series of 42 influenza infected postpartum mothers taking precautions (hand hygiene and wearing masks) before breastfeeding showed that no neonates were infected with influenza during one-month of follow-up.

## Author contributions

(I) Conception and design: E Liu, Y Chen; (II) Administrative support: None; (III) Provision of study materials or patients: N Yang, S Che; (IV) Collection and assembly of data: Nan Yang, J Zhang, Xia Wang; (V) Data analysis and interpretation: N Yang, S Che, Y Tang; (VI) Manuscript writing: All authors; (VII) Final approval of manuscript: All authors.

## Acknowledgments

We thank Janne Estill, Institute of Global Health of University of Geneva for providing guidance and comments for our review. We thank all the authors for their wonderful collaboration.

## Funding

This work was supported by grants from National Clinical Research Center for Child Health and Disorders (Children’s Hospital of Chongqing Medical University, Chongqing, China) (grant number NCRCCHD-2020-EP-01) to [Enmei Liu]; Special Fund for Key Research and Development Projects in Gansu Province in 2020, to [Yaolong Chen]; The fourth batch of “Special Project of Science and Technology for Emergency Response to COVID-19” of Chongqing Science and Technology Bureau, to [Enmei Liu]; Special funding for prevention and control of emergency of COVID-19 from Key Laboratory of Evidence Based Medicine and Knowledge Translation of Gansu Province (grant number No. GSEBMKT-2020YJ01), to [Yaolong Chen].

## Footnote

### Conflicts of Interest

The authors have no conflicts of interest to declare.

### Ethical Statement

The authors are accountable for all aspects of the work in ensuring that questions related to the accuracy or integrity of any part of the work are appropriately investigated and resolved.

## Supplementary Material 1-Search strategy

### PubMed

#1. “COVID-19” [Supplementary Concept]

#2. “severe acute respiratory syndrome coronavirus 2”[Supplementary Concept]

#3. “Novel coronavirus” [Title/Abstract]

#4. “2019-novel coronavirus”[Title/Abstract]

#5. “Novel CoV”[Title/Abstract]

#6. “2019-nCoV”[Title/Abstract]

#7. “2019-CoV”[Title/Abstract]

#8. “Wuhan-Cov”[Title/Abstract]

#9. “Wuhan Coronavirus”[Title/Abstract]

#10. “Wuhan seafood market pneumonia virus”[Title/Abstract]

#11. “Middle East Respiratory Syndrome Coronavirus”[Mesh]

#12. “Middle East Respiratory Syndrome”[Title/Abstract]

#13. “MERS” [Title/Abstract]

#14. “MERS-CoV” [Title/Abstract]

#15. “Severe Acute Respiratory Syndrome”[Mesh]

#16. “SARS Virus”[Mesh]

#17. “Severe Acute Respiratory Syndrome”[Title/Abstract]

#18. “SARS”[Title/Abstract]

#19. “SARS-CoV”[Title/Abstract]

#20. “SARS-Related” [Title/Abstract]

#21. “SARS-Associated”[Title/Abstract]

#22. “SARS-CoV-2”[Title/Abstract]

#23. “COVID-19”[Title/Abstract]

#24. “Influenzavirus A”[Mesh]

#25. “Influenzavirus B”[Mesh]

#26. “Influenza, Human”[Mesh]

#27. “Influenzavirus A”[Title/Abstract]

#28. “Influenzavirus B”[Title/Abstract]

#29. Influenza*[Title/Abstract]

#30. flu[Title/Abstract])

#31. h1n1[Title/Abstract])

#32. h5n1[Title/Abstract]

#33. h3n2[Title/Abstract]

#34. #1-33/ OR

#35. “Breast Feeding”[Mesh]

#36. “Lactation”[Mesh]

#37. “breastfe*”[Title/Abstract]

#38. “breast-fe*”[Title/Abstract]

#39. “breast fe*”[Title/Abstract]

#40. “lactation*”[Title/Abstract]

#41. milk[Title/Abstract]

#42. “infant fe*”[Title/Abstract]

#43. “baby fe*”[Title/Abstract]

#44. #35-43/ OR

#45. #34 AND #44

### Embase

#1. ‘novel coronavirus’:ab,ti

#2. ‘2019-novel coronavirus’:ab,ti

#3. ‘novel cov’:ab,ti

#4. ‘2019-ncov’:ab,ti

#5. ‘2019-cov’:ab,ti

#6. ‘wuhan-cov’:ab,ti

#7. ‘wuhan coronavirus’:ab,ti

#8. ‘wuhan seafood market pneumonia virus’:ab,ti

#9. ‘middle east respiratory syndrome coronavirus’/exp

#10. ‘middle east respiratory syndrome’:ab,ti

#11. ‘middle east respiratory syndrome coronavirus’:ab,ti

#12. ‘mers’:ab,ti

#13. ‘mers-cov’:ab,ti

#14. ‘severe acute respiratory syndrome’/exp

#15. ‘sars coronavirus’/exp

#16. ‘severe acute respiratory syndrome’:ab,ti

#17. ‘sars’:ab,ti

#18. ‘sars-cov’:ab,ti

#19. ‘sars-related’:ab,ti

#20. ‘sars-associated’:ab,ti

#21. ‘sars-cov-2’:ab,ti

#22. ‘covid-19’:ab,ti

#23. h1n1:ab,ti

#24. h5n1:ab,ti

#25. h3n2:ab,ti

#26. influenza*:ab,ti

#27. flu:ab,ti

#28. ‘influenza virus a’/exp

#29. ‘influenza virus b’/de

#30. ‘influenza’/exp

#31. #1-30/ OR

#32. ‘breast feeding’/exp

#33. ‘lactation’/exp

#34. ‘breastfe*’:ab,ti

#35. ‘breast-fe*’:ab,ti

#36. ‘breast fe*’:ab,ti

#37. ‘lactation*’:ab,ti

#38. ‘milk’:ab,ti

#39. ‘infant fe*’:ab,ti

#40. ‘baby fe*’:ab,ti

#41. #32-40/ OR

#42. #31 AND #41

### Cochrane library

#1. “Novel coronavirus”:ti,ab,kw

#2. “2019-novel coronavirus”:ti,ab,kw

#3. “Novel CoV”:ti,ab,kw

#4. “2019-nCoV”:ti,ab,kw

#5. “2019-CoV”:ti,ab,kw

#6. “Wuhan-Cov”:ti,ab,kw

#7. “Wuhan Coronavirus”:ti,ab,kw

#8. “Wuhan seafood market pneumonia virus”:ti,ab,kw

#9. MeSH descriptor: [Middle East Respiratory Syndrome Coronavirus] explode all trees

#10. “Middle East Respiratory Syndrome”:ti,ab,kw

#11. “MERS”:ti,ab,kw

#12. “MERS-CoV”:ti,ab,kw

#13. MeSH descriptor: [Severe Acute Respiratory Syndrome] explode all trees

#14. MeSH descriptor: [SARS Virus] explode all trees

#15. “Severe Acute Respiratory Syndrome”:ti,ab,kw

#16. “SARS”:ti,ab,kw

#17. “SARS-CoV”:ti,ab,kw

#18. “SARS-Related”:ti,ab,kw

#19. “SARS-Associated”:ti,ab,kw

#20. “sars-cov-2”:ti,ab,kw

#21. “covid-19”:ti,ab,kw

#22. MeSH descriptor: [Influenza, Human] explode all trees

#23. MeSH descriptor: [Influenzavirus A] explode all trees

#24. MeSH descriptor: [Influenzavirus B] explode all trees

#25. “Influenzavirus B”:ti,ab,kw

#26. “Influenzavirus A”:ti,ab,kw

#27. “Influenza*”:ti,ab,kw

#28. “flu”:ti,ab,kw

#29. “h1n1”:ti,ab,kw

#30. “h5n1”:ti,ab,kw

#31. “h3n2”:ti,ab,kw

#32. #1-31/ OR

#33. MeSH descriptor: [breastfeeding] explode all trees

#34. MeSH descriptor: [lactation] explode all trees

#35. “breastfe*”:ti,ab,kw

#36. “breast-fe*”:ti,ab,kw

#37. “breast fe*”:ti,ab,kw

#38. “lactation*”:ti,ab,kw

#39. “milk”:ti,ab,kw

#40. “infant fe*”:ti,ab,kw

#41. “baby fe*”:ti,ab,kw

#42. #33-41/OR

#43. #32 AND #42

### Web of Science

#1. TOPIC: “Novel coronavirus”

#2. TOPIC: “2019-novel coronavirus”

#3. TOPIC: “Novel CoV”

#4. TOPIC: “2019-nCoV”

#5. TOPIC: “2019-CoV”

#6. TOPIC: “Wuhan-Cov”

#7. TOPIC: “Wuhan Coronavirus”

#8. TOPIC: “Wuhan seafood market pneumonia virus”

#9. TOPIC: “Middle East Respiratory Syndrome”

#10. TOPIC: “MERS”

#11. TOPIC: “MERS-CoV”

#12. TOPIC: “Severe Acute Respiratory Syndrome”

#13. TOPIC: “SARS”

#14. TOPIC: “SARS-CoV”

#15. TOPIC: “SARS-Related”

#16. TOPIC: “SARS-Associated”

#17. TOPIC: “SARS-CoV-2”

#18. TOPIC: “COVID-19”

#19. TOPIC: “Influenzavirus B”

#20. TOPIC: “Influenzavirus A”

#21. TOPIC: “Influenza*”

#22. TOPIC: “flu”

#23. TOPIC: “h1n1”

#24. TOPIC: “h5n1”

#25. TOPIC: “h3n2”

#26. #1-25/ OR

#27. TOPIC: “breastfe*”

#28. TOPIC: “breast-fe*”

#29. TOPIC: “breast fe*”

#30. TOPIC: “lactation*”

#31. TOPIC: “milk”

#32. TOPIC: “infant fe*”

#33. TOPIC: “baby fe*”

#34. #27-33/ OR

#35. #26 AND #34

### CBM

**#1**. “新型冠状病毒”[常用字段:智能]

**#2**. “2019-nCoV”[常用字段:智能]

**#3**. “2019-CoV”[常用字段:智能]

**#4**. “武汉冠状病毒”[5E38用字段:智能]

**#5**. “中东呼吸综合征冠状病毒”[不加权:扩展]

**#6**. “中东呼吸综合征”[常用字段:智能]

**#7**. “MERS”[常用字段:智能]

**#8**. “MERS-CoV”[常用字段:智能]

**#9**. “严重急性呼吸综合征”[不加权:扩展]

**#10**. “SARS 病毒”[不加权:扩展]

**#11**. “严重急性呼吸综合征”

**#12**. “SARS”[常用字段:智能]

**#13**. “非典型肺炎”[常用字段:智能]

**#14**. “SARS-CoV-2”[常用字段:智能]

**#15**. “COVID-19”[常用字段:智能]

**#16**. “甲流”[常用字段:智能]

**#17**. “乙流”[常用字段:智能]

**#18**. “流行性感冒”[常用字段:智能]

**#19**. “流感”[常用字段:智能]

**#20**. “H1N1”[常用字段:智能]

**#21**. “H5N1”[常用字段:智能]

**#22**. “H3N2”[常用字段:智能]

**#23**. “流感, 人”[不加权:扩展]

**#24**. “流感病毒属A 型”[不加权:扩展]

**#25**. “流感病毒属B 型”[不加权:扩展]

**#26**. #1-25/ OR

**#27**. “母乳喂养”[不加权:扩展]

**#28**. “乳, 人”[不加权:扩展] OR

**#29**. “;乳”[常用字段:智能]

**#30**. “喂”[常用字段:智能]

**#31**. “奶”[常用字段:智能]

**#32**. #27-31/ OR

**#33**. #26 AND #32

### WanFang

**#1**. “新型冠状病毒”[主题]

**#2**. “2019-nCoV”[主题]

**#3**. “2019-CoV”[主题]

**#4**. “武汉冠状病毒”[主题]

**#5**. “中东呼吸综合征”[主题]

**#6**. “MERS”[主题]

**#7**. “MERS-CoV”[主题]

**#8**. “严重急性呼吸综合征”[主题]

**#9**. “SARS”[主题]

**#10**. “非典型肺炎”[主题]

**#11**. “SARS-CoV-2”[主题]

**#12**. “COVID-19”[主题]

**#13**. “甲流”[主题]

**#14**. “乙流”[主题]

**#15**. “流行性感冒”[主题]

**#16**. “流感”[主题]

**#17**. “H1N1”[主题]

**#18**. “H5N1”[主题]

**#19**. “H3N2”[主题]

**#20**. #1-19/ OR

**#21**. “乳”[主题]

**#22**. “喂”[主题]

**#23**. “奶”[主题]

**#24**. #21-23/ OR

**#25**. #20 AND #24

### CNKI

**#1**. “新型冠状病毒”[主题]

**#2**. “2019-nCoV”[主题]

**#3**. “2019-CoV”[主题]

**#4**. “武汉冠状病毒”[主题]

**#5**. “中东呼吸综合征”[主题]

**#6**. “MERS”[主题]

**#7**. “MERS-CoV”[主题]

**#8**. “严重急性呼吸综合征”[主题]

**#9**. “SARS”[主题]

**#10**. “非典型肺炎”[主题]

**#11**. “SARS-CoV-2”[主题]

**#12**. “COVID-19”[主题]

**#13**. “甲流”[主题]

**#14**. “乙流”[主题]

**#15**. “流行性感冒”[主题]

**#16**. “H1N1”[主题]

**#17**. “H5N1”[主题]

**#18**. “H3N2”[主题]

**#19**. #1-18/ OR

**#20**. “乳”[主题]

**#21**. “喂”[主题]

**#22**. “奶”[主题]

**#23**. #20-22/ OR

**#24**. #19 AND #23

## Notes

### Competing Interest Statement

The authors have declared no competing interest.

